# Prevalence of Intestinal Parasitic Infections Among HIV-Infected Patients: A Five-Year Retrospective Study

**DOI:** 10.1101/2025.06.01.25328736

**Authors:** Duressa Shafi Ahmed

## Abstract

**Background:** Intestinal parasitic infections remain a significant health concern for individuals living with HIV/AIDS, particularly in resource-limited settings such as Ethiopia. Due to compromised immune function, HIV-infected patients are highly susceptible to secondary infections, including parasitic diseases, which can exacerbate their health conditions. Understanding the prevalence and distribution of these infections is crucial for improving patient care and intervention strategies.

**Method:** A retrospective cross-sectional study was conducted to assess the prevalence of intestinal parasites among HIV-infected patients attending Hossana Health Center over the past five years. The study utilized a convenience sampling method, including data from all patients recorded in the facility’s logbook. The completeness of patient records was verified before data extraction. Chi-square tests were performed using R programming software to examine associations between demographic variables and parasitic infections. A p-value of less than 0.05 was considered statistically significant, and results were presented in frequency tables for clarity.

**Results:** The analysis revealed an overall prevalence of intestinal parasitic infections at 31.93% among HIV-infected patients. The most frequently identified parasites included *Ascaris lumbricoides* (42.47%), *Entamoeba histolytica* (27.82%), and *Giardia lamblia* (9.20%). Other detected parasites included *Trichuris trichiura* (5.86%) and *Taenia species* (5.02%), which were the least prevalent. A statistically significant association was observed between age groups and parasitic infection rates (p < 0.001), suggesting that age-related factors—such as environmental exposure, behavioral patterns, and differences in immune function—may contribute to the likelihood of infection.

**Conclusion:** The findings highlight the persistent burden of intestinal parasitic infections among HIV-infected individuals and reinforce the importance of age-specific intervention strategies. Efforts should focus on improving sanitation, hygiene, and targeted health education programs to mitigate the impact of these infections in vulnerable populations.

## Introduction

Intestinal parasitic infections (IPIs) remain a major global health concern, particularly in resource-limited settings, where they contribute significantly to morbidity and mortality. It is estimated that over three billion people worldwide are infected with one or more intestinal parasites, disproportionately affecting populations with limited access to clean water, sanitation, and healthcare services (1). These infections pose an even greater challenge for immunocompromised individuals, particularly HIV/AIDS patients, due to the complex interaction between parasitic pathogens and immune function (2).

The link between IPIs and HIV/AIDS progression is well-documented. Chronic parasitic infections stimulate immune activation, leading to rapid CD4 cell depletion, which accelerates HIV progression (3). Conversely, HIV-induced immunosuppression makes individuals more vulnerable to both opportunistic and non-opportunistic parasitic infections, further exacerbating gastrointestinal complications (4).

Sub-Saharan Africa bears a disproportionate burden of the HIV/AIDS epidemic, accounting for approximately 67% of global HIV cases (5). In Ethiopia, where poor sanitation and limited healthcare access persist, HIV/AIDS is further compounded by a high prevalence of intestinal parasitic infections (2).

Studies have reported that gastrointestinal complications—particularly chronic diarrhea— occur in 50–96% of HIV/AIDS cases globally, with prevalence rates reaching 90% in parts of Africa (3). Although antiretroviral therapy (ART) has reduced these infections in some regions, limited ART access in resource-poor settings has allowed parasitic infections to remain widespread, severe, and frequently recurrent (6).

Several opportunistic protozoan pathogens, including *Isospora belli*, Cryptosporidium parvum, *Cyclospora cayetanensis*, and *Microsporidia species*, are frequently implicated in gastrointestinal disease in HIV/AIDS patients (7,8). Additionally, helminth infections contribute to immune dysregulation, worsening health outcomes. Research indicates that HIV viral loads are significantly higher in co-infected individuals, while anti-parasitic treatments correlate with viral load reductions (9,10).

Since the emergence of HIV/AIDS, co-infection with intestinal parasites has remained a persistent global challenge (11). In Ethiopia, approximately 613,000 individuals live with HIV, and the burden of new infections and related complications remains substantial (12).

Given the interconnected nature of HIV/AIDS and intestinal parasitic infections, comprehensive epidemiological studies are essential to inform targeted public health interventions. This study aims to contribute to that knowledge by assessing the prevalence of intestinal parasitic infections among HIV-infected patients based on a five-year retrospective review of clinical records at Hossana Health Center.

## Methods

### Study Design and Study Area

This retrospective cross-sectional study was conducted at Hossana Health Center, located in Hossana Town, approximately 231 km south of Addis Ababa, Ethiopia. The town sits at an altitude of 2,177 meters above sea level, with temperatures ranging from 13°C at night to 24°C during the day, and an annual rainfall between 1,500 and 1,800 mm. Based on 2020 population estimates, Hossana had 75,963 residents.

The data review took place from December 2 to 28, 2020, covering medical records from January 2016 to December 2020. Data were extracted from patient logbooks maintained at Hossana Health Center to assess the prevalence of intestinal parasitic infections among HIV/AIDS patients.

### Sampling Technique and Sample Size

A convenience sampling technique was employed, including all HIV-infected patients with complete stool examination records in the logbooks during the study period. The final sample consisted of 1,497 patient records spanning the five-year retrospective review.

### Data Collection Process

Patient data were retrieved from logbooks at Hossana Health Center using a standardized data collection form to systematically record relevant variables, including age, sex, and stool examination results. Trained data collectors ensured accuracy in the extraction process, with the information directly digitized for analysis after being recorded.

### Data Processing and Analysis

After extracting patient information from logbook records at Hossana Health Center, the data were systematically recorded and organized before being entered into R programming for analysis. Descriptive statistics were used to summarize socio-demographic characteristics and the prevalence of intestinal parasitic infections. To assess the relationship between the dependent variable (intestinal parasitic infection) and independent variables (age and sex), a chi-square test was performed, with statistical significance set at p < 0.05.

### Ethical Approval and Considerations

Ethical approval for this study was obtained from the Institutional Review Board (IRB) of the College of Medicine and Health Sciences at Wachemo University in 2020. This research relied exclusively on retrospective data from logbooks at Hossana Health Center, with no direct contact with patients. As a result, individual informed consent was not required. All procedures adhered to relevant ethical guidelines and regulations.

To ensure ethical integrity, the following measures were taken:

- A letter of permission was obtained from Wachemo University and the appropriate health authorities for access to the logbook data.
- All data were handled with strict confidentiality, ensuring that no identifiable patient information was accessed or disclosed.
- The study was conducted without disrupting ongoing health programs or patient care, as it focused solely on historical, non-invasive records.

## Result

### Socio-demography of the study subjects

A total of 1497 individuals were screened for intestinal parasite during the study period. Out of these participants 51.64% were female and 48.36 % were male (Table 1). Concerning the study participants, 94.52% were found between 15-60 age groups, 0.94% were less than 15 age groups and 4.54% were above 60 age groups (Table 2).

**Table 1.**
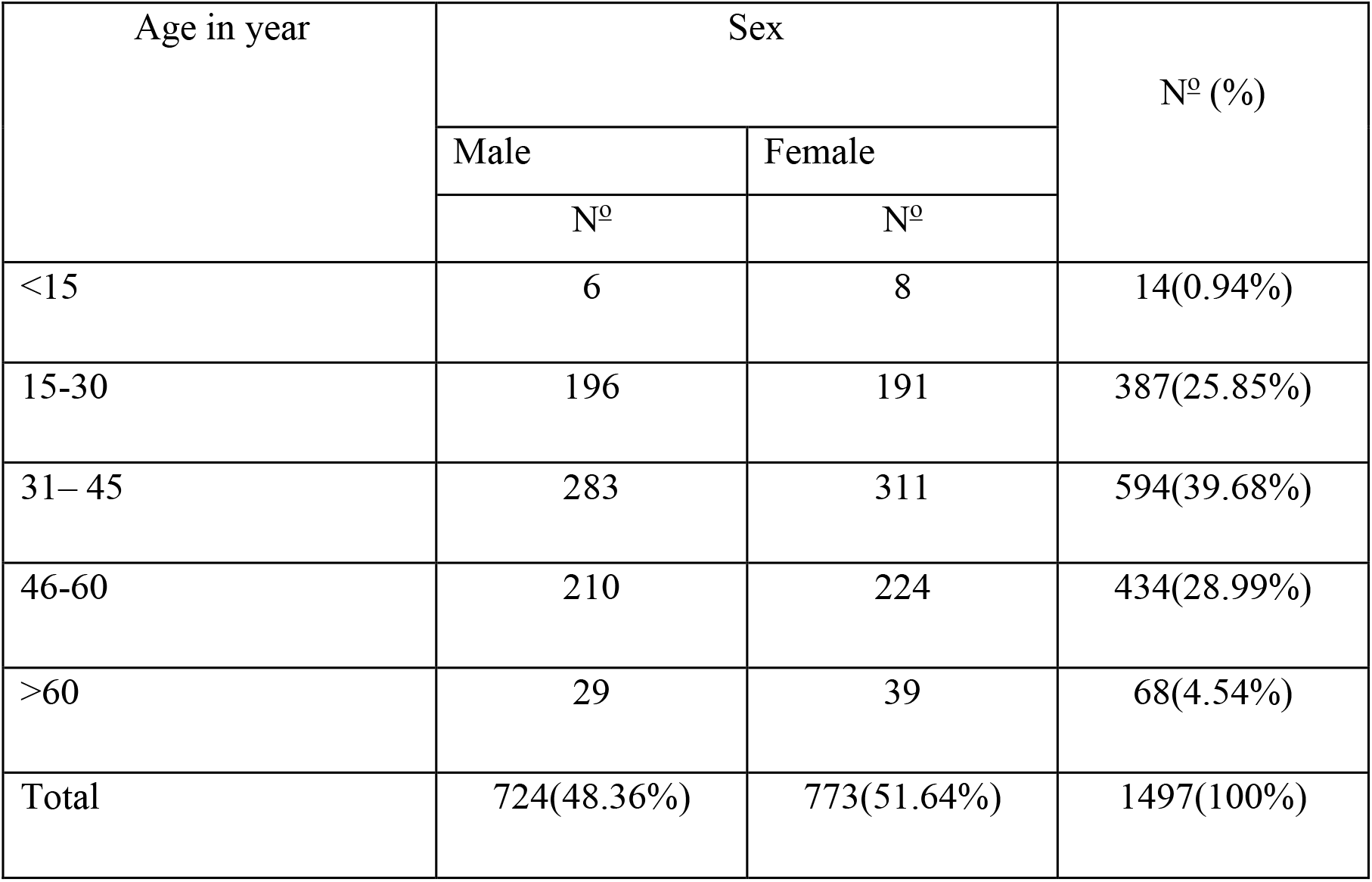
Socio-demographic characteristics among HIV infected patients from January 2016 - December 2020 at Hossana Health center.

**Table 2.**
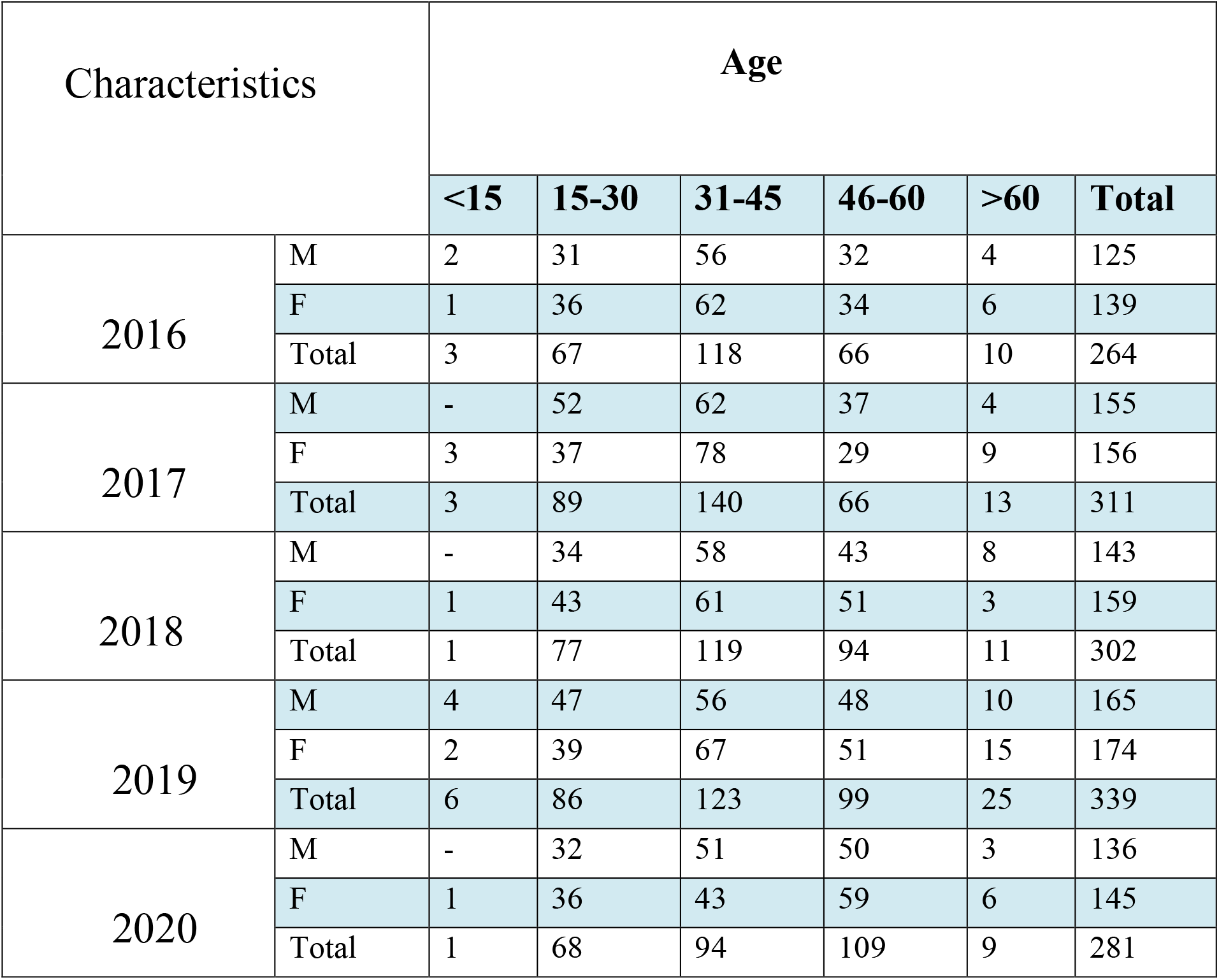
Socio-demographic characteristics among HIV infected patients from January 2016 - December 2020 at Hossana Health center.

| Characteristics |  | Age |  |  |  |  |  |
| --- | --- | --- | --- | --- | --- | --- | --- |
|  |  | <15 | 15-30 | 31-45 | 46-60 | >60 | Total |
| 2016 | M | 2 | 31 | 56 | 32 | 4 | 125 |
|  | F | 1 | 36 | 62 | 34 | 6 | 139 |
|  | Total | 3 | 67 | 118 | 66 | 10 | 264 |
| 2017 | M | - | 52 | 62 | 37 | 4 | 155 |
|  | F | 3 | 37 | 78 | 29 | 9 | 156 |
|  | Total | 3 | 89 | 140 | 66 | 13 | 311 |
| 2018 | M | - | 34 | 58 | 43 | 8 | 143 |
|  | F | 1 | 43 | 61 | 51 | 3 | 159 |
|  | Total | 1 | 77 | 119 | 94 | 11 | 302 |
| 2019 | M | 4 | 47 | 56 | 48 | 10 | 165 |
|  | F | 2 | 39 | 67 | 51 | 15 | 174 |
|  | Total | 6 | 86 | 123 | 99 | 25 | 339 |
| 2020 | M | - | 32 | 51 | 50 | 3 | 136 |
|  | F | 1 | 36 | 43 | 59 | 6 | 145 |
|  | Total | 1 | 68 | 94 | 109 | 9 | 281 |

### Prevalence of intestinal parasites

There were about 478(31.93%) patients positive for one or more intestinal parasites (Table 3). The most frequently detected parasites *were Ascaris lumbricoides* 203(42.47%) followed by *Entamoeba histolytica* 133(27.82%) and *Giardia lamblia* 44(9.2%) (Table 4 & Figure 1). The 16– 30 and 31–45 age groups showed the highest parasite infections across all years. *Ascaris lumbricoides* was the most frequently detected parasite among both sexes (Table 5).

**Table 3.** Prevalence of intestinal parasites among HIV infected patients (2016-2020) at HHC.

| Year | N<sub>0</sub> of stool examination among HIV patients | N<sub>0</sub> of positive intestinal parasite results |  |
| --- | --- | --- | --- |
|  |  | By Number | By percentage (%) |
| 2016 | 264 | 103 | 39.02 |
| 2017 | 311 | 128 | 41.16 |
| 2018 | 302 | 94 | 31.13 |
| 2019 | 339 | 115 | 33.92 |
| 2020 | 281 | 38 | 20.99 |
| **Total** | 1497 | 478 | 31.93 |

**Table 4.** Prevalence of each intestinal parasite species among HIV infected patients (2016-2020) at HHC.

| Year | Total N<sub>0</sub> of each intestinal parasite species |  |  |  |  |  |
| --- | --- | --- | --- | --- | --- | --- |
|  | A. lumbricoides | H. worm | T. species | T. trichiura | G. lamblia | E. histolytica |
| 2016 | 49 | 8 | 4 | 6 | 12 | 23 |
| 2017 | 64 | 8 | 6 | 9 | 14 | 26 |
| 2018 | 33 | 11 | 4 | 3 | 6 | 36 |
| 2019 | 43 | 10 | 8 | 7 | 9 | 37 |
| 2020 | 14 | 5 | 2 | 3 | 3 | 11 |
| **Total** | 203 | 42 | 24 | 28 | 44 | 133 |

**Figure 1.**
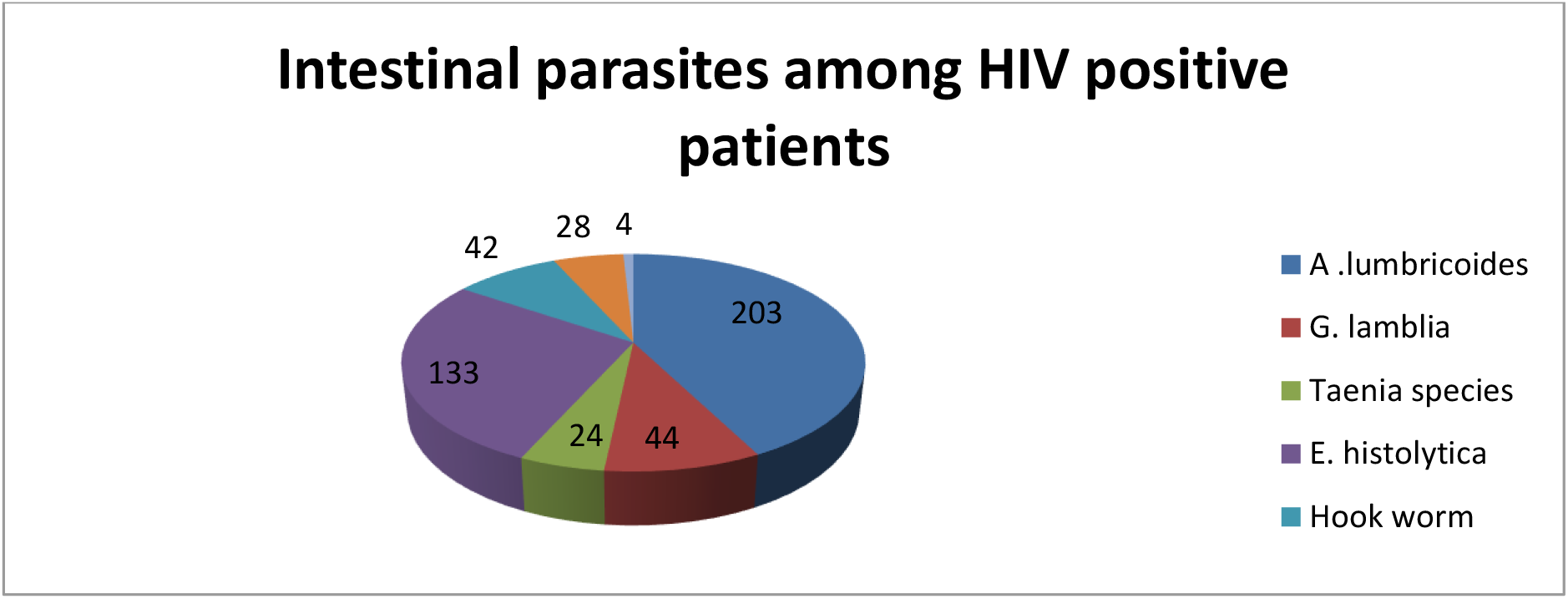
Percentage distribution of parasitic infection among HIV infected patients (2016-2020) at HHC.

**Table 5.**
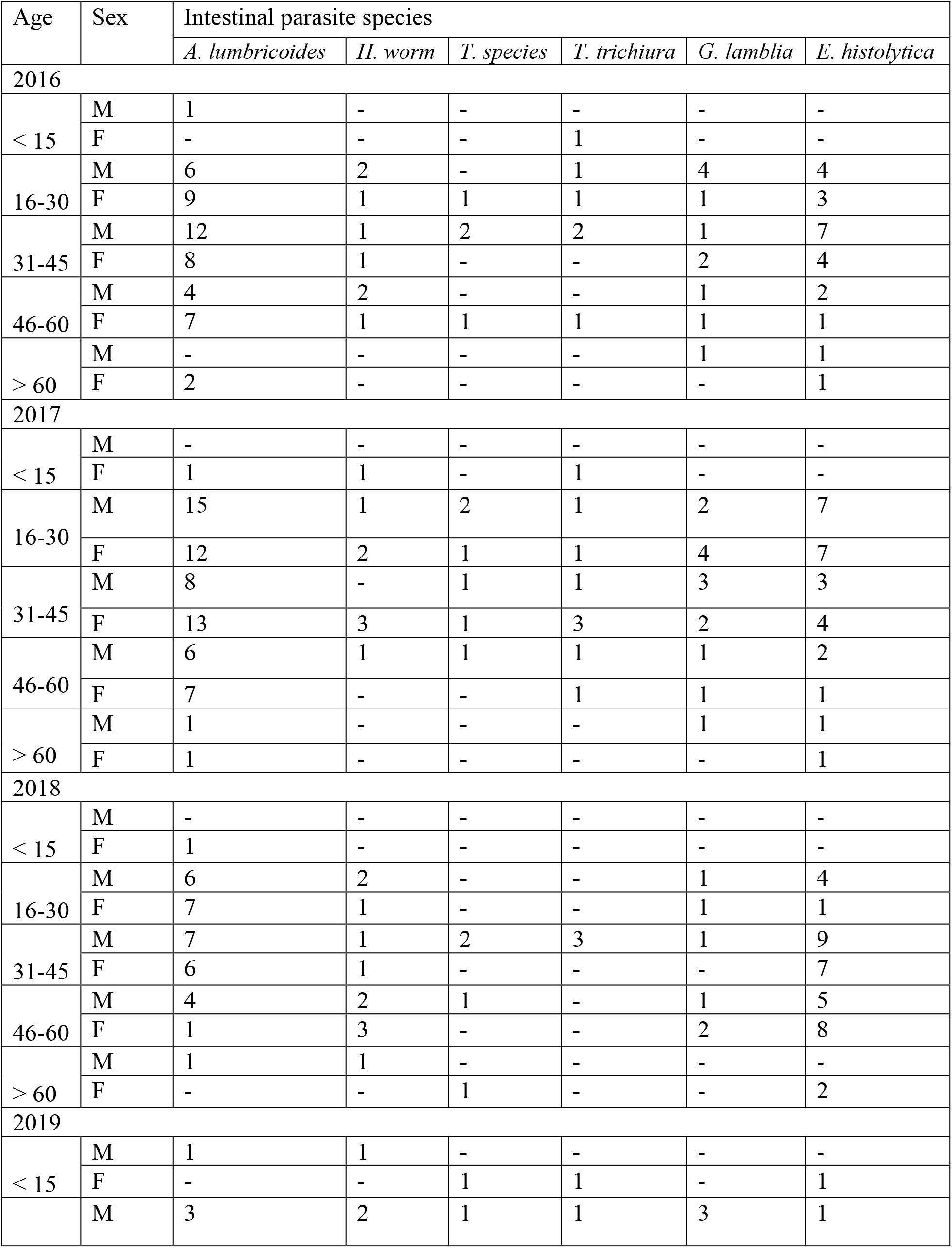
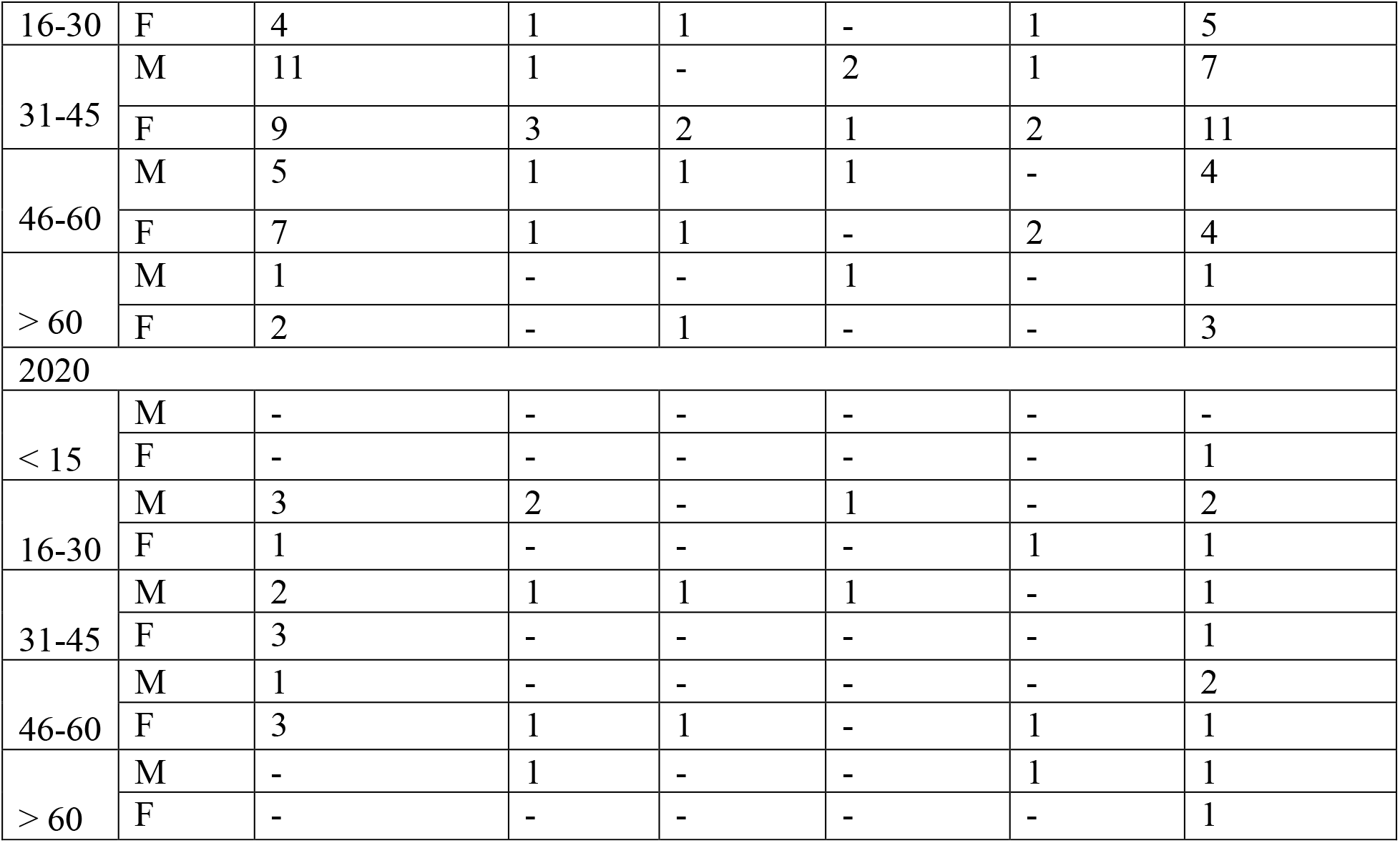
Age and Sex Distribution of Intestinal Parasite Infections Among HIV-Infected Patients (2016*-*2020) at H*HC*.

### Chi-Square result of Intestinal Parasitic Infections by Sex and Age Group Among HIV-Infected Patients

The chi-square test results indicate no statistically significant relationship between sex and the prevalence of intestinal parasitic infections (χ2 = 0.47, p ≈ 0.50). Male and female participants had similar proportions of positive cases. In contrast, the chi-square test for age groups revealed a highly significant association (χ2 = 38.36, p < 0.001), indicating that age significantly affects the prevalence of intestinal parasitic infections. The 31–45 age group had the highest number of positive cases (179 patients), while the **<**15 age group had the lowest (12 patients) (table 6).

**Table 6.**
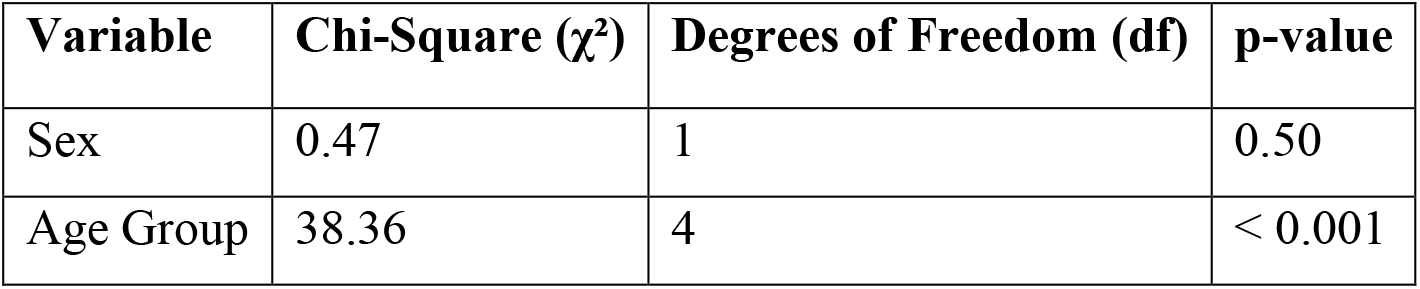
Chi-Square result of Intestinal Parasitic Infections by Sex and Age Group Among HIV-Infected Patients.

| Variable | Chi-Square ($\chi^2$) | Degrees of Freedom (df) | p-value |
| --- | --- | --- | --- |
| Sex | 0.47 | 1 | 0.50 |
| Age Group | 38.36 | 4 | < 0.001 |

## Discussions

The present five-year retrospective study at Hossana Health Center revealed that 31.93% of HIV-infected patients had one or more intestinal parasitic infections. This finding is in line with recent Ethiopian studies showing prevalence rates ranging from approximately 21.1% to 35.1% among similar populations (13,14). The relatively high burden of intestinal parasites, particularly in a resource-limited setting, underscores the continuing public health challenge these infections pose for immunocompromised individuals.

Our study demonstrated that the overall prevalence of intestinal parasitic infections was 31.93%, with *Ascaris lumbricoides* and *Entamoeba histolytica* being the most frequently detected species (42.47% and 27.82% of positive cases, respectively). These figures are comparable to other recent reports from Ethiopia where modest to high infection rates have been attributed to factors such as poor sanitation, low socioeconomic status, and inadequate hygiene practices (13–16). The high proportion of *A. lumbricoides* could be associated with environmental contamination and suboptimal handwashing practices, while the prominence of *E. histolytica* may reflect challenges in water quality and personal hygiene (16).

The majority of the study population (94.52%) were between the ages of 15 and 60 years, which corresponds to the most economically active group. Notably, the highest infection rates were observed in the 31–45 years age group in several years. This is particularly concerning because intestinal parasites can lead not only to chronic diarrhea and nutritional deficiencies (17,18), but also to impaired work capacity and reduced quality of life in this key demographic (19,20). Future interventions should target these age groups with tailored health education and improved access to sanitation facilities.

Our data show variability in prevalence over the study period. For instance, the prevalence peaked at 41.16% in 2017 and dropped to 20.99% in 2020. While this fluctuation might partly reflect improvements in public health interventions, such as enhanced water, sanitation, and hygiene (WASH) initiatives (21), it could also be influenced by changes in diagnostic practices or sample sizes (22). Further prospective studies are necessary to ascertain whether these differences represent a true change in the epidemiology or are artifacts of study design.

When compared with data from other regions, our overall prevalence is similar to figures documented in Cameroon (27.9%) (23), although it remains lower than those reported in some studies from Thailand and other parts of Ethiopia (up to 50%) (24). Such differences likely emanate from geographical, environmental, and socioeconomic variations, as well as differences in the performance of the diagnostic techniques. Studies using more sensitive methods (e.g., molecular assays) often report higher detection rates (25).

The relationship between intestinal parasites and HIV is bidirectional. Intestinal parasites can contribute to chronic immune activation and skew the immune response toward a T-helper-2 profile, potentially accelerating HIV progression (26). Conversely, the immunosuppressive state induced by HIV infection increases susceptibility to both opportunistic and non-opportunistic parasitic infections (27,28). Recent evidence indicates that co-infection with helminths is associated with higher HIV viral loads, and that effective anti-parasitic treatment may reduce this viral burden (29). This intertwined relationship makes the diagnosis and management of intestinal parasites a crucial component of comprehensive HIV care (30).

The final consolidated table succinctly presents the results of the chi-square tests performed on demographic variables. The analysis indicates that while there is no statistically significant correlation between sex and the prevalence of intestinal parasitic infections (p = 0.50), the association with age groups is highly significant (p < 0.001). This suggests that age-related factors—such as behavioral patterns, environmental exposure, and biological differences in immune function—may influence the likelihood of parasitic infection among HIV-infected individuals, whereas sex does not appear to be a determining factor (27,31).

These findings highlight the necessity of age-specific public health interventions and further research into age-related disparities in exposure and susceptibility (13,27). Implementing targeted prevention and treatment strategies for high-risk age groups could significantly improve the management of intestinal parasitic infections among HIV-infected populations in resource-limited settings (13,29).

## Limitations

The retrospective nature of this study inherently limits the interpretation of causality. Additionally, reliance on conventional stool examination methods might underestimate the true prevalence because of their lower sensitivity compared with advanced techniques such as polymerase chain reaction (PCR). Lack of detailed data on patients’ antiretroviral therapy (ART) status and immune parameters (like CD4 counts) further limits the ability to correlate parasitic infections with the degree of immunosuppression.

## Conclusion

This study highlights the substantial burden of intestinal parasitic infections among HIV-infected patients at Hossana Health Center. The findings emphasize the urgent need for integrated diagnostic, therapeutic, and preventive strategies tailored to resource-limited settings. Strengthening public health infrastructure and incorporating routine screening for parasitic infections into HIV care services may help reduce morbidity and improve overall health outcomes for HIV-infected individuals.

Given the high prevalence of specific parasites such as *Ascaris lumbricoides* and *Entamoeba histolytica*, comprehensive public health interventions are essential. Enhancing diagnostic approaches, particularly by integrating sensitive tools like PCR-based assays alongside traditional microscopy, can improve detection and characterization of parasitic infections. Additionally, implementing WASH (Water, Sanitation, and Hygiene) programs and promoting behavioral changes—especially in high-risk age groups—will play a crucial role in reducing infections. Routine deworming strategies within HIV care settings should be prioritized.

Prospective cohort studies are necessary to explore the dynamic relationship between parasitic co-infection and HIV progression, particularly in relation to viral load and CD4 count fluctuations over time. These studies can provide valuable insights into disease management and inform effective treatment protocols.

Policy implementation is another critical area that requires attention. Policymakers should incorporate routine parasitic screening into HIV management protocols while improving infrastructural measures that ensure access to safe water and adequate sanitation facilities. Furthermore, community education campaigns focused on hand hygiene, proper food preparation, and the risks associated with poor sanitation should be expanded, particularly in regions with high HIV prevalence.

By addressing these challenges with a multi-faceted approach, health authorities and researchers can work toward mitigating the impact of intestinal parasitic infections among HIV-infected populations, ultimately improving health outcomes and quality of life.

## Data Availability

All data produced in the present study are available upon reasonable request to the authors

## Abbreviations

AIDS: Acquired Immuno-deficiency Syndrome
ART: Anti-Retroviral Treatment
CD4: Cluster for Differentiation
CNS: Central Nervous System
HIV: Human Immunodeficiency Virus
IP: Intestinal Parasite
MOH: Ministry of Health
UNAIDS: United Nation Program on HIV/AIDS
HHC: Hossana Health Center

## Acknowledgments

We extend our sincere appreciation to the dedicated data collectors and laboratory staff at the Hossana Health Center for their invaluable contributions to this study. Additionally, we are grateful to all the staff members of HHC for their support, particularly in assisting with data retrieval from the logbooks. Their efforts have been instrumental in ensuring the accuracy and completeness of this research.

## Ethical Considerations

Ethical issues were carefully considered throughout the study. Key measures included:

1. Obtaining a formal letter of permission from Wachemo University and the appropriate health authorities.
2. Ensuring that all information obtained during the course of this investigation was kept confidential.
3. Conducting the study in a manner that did not interfere with ongoing health programs or affect patient care, as this study involved a review of non-invasive, secondary data only.

## Availability of data and materials

The data set used can be obtained from the corresponding author.

## Authors’ contributions

### DSA

Proposal development, data retrieval from the logbooks., data analysis, drafting and final preparation of the manuscript.

### Consent for publication

Not applicable

